# Antibody boosting and short-term durability following SARS-CoV-2 infections in United States longitudinal cohorts, 2020–2024

**DOI:** 10.64898/2026.09.17.26363304

**Authors:** Jade Yangyupei Yang, Anastasiya Risukhina, Amy Callear, Casey Juntila-Raymond, Emileigh Johnson, Elie-Tino Godonou, Matthew Smith, Claire M. Midgley, Jefferson M. Jones, Arnold S. Monto, Emily T. Martin

**Author notes:** Corresponding author: Emily T. Martin.

## Abstract

To characterize anti-nucleocapsid (anti-N) IgG binding antibody responses following first PCR-confirmed SARS-CoV-2 infections in a highly vaccinated population, we evaluated antibody boosting and durability in a longitudinal cohort in Michigan. Among 113 participants with paired pre- and post-infection serum specimens tested using a multiplex electrochemiluminescence assay, SARS-CoV-2 infection induced substantial boosting of anti-N IgG concentrations; approximately 80% of participants becoming anti-N seropositive following infection, and 81% of participants demonstrating at least a 4-fold rise in antibody concentrations following infection. Population-level analyses of 66 participants with post-infection serum specimens collected 52–120 days after infection demonstrated sustained anti-N IgG concentrations over time, with no evidence of substantial decline. These findings demonstrate robust boosting and sustained infection-induced anti-N IgG responses in a highly vaccinated population through four months following SARS-CoV-2 infection. This supports the use of anti-N antibodies for longitudinal serosurveillance, including those who are vaccinated.

## Introduction

Current U.S.-approved COVID-19 vaccines target the spike protein of SARS-CoV-2, not the nucleocapsid (N) protein^1,2^. As a result, antibodies against the nucleocapsid (anti-N) protein are generally produced only after infection, making them an important marker for infection-based serosurveillance. Some prior research suggested that anti-N IgG antibody response may be attenuated in individuals vaccinated before infection, raising questions about the strength, durability and utility of an anti-N IgG marker in highly vaccinated populations^3–5^. However, other studies have found that vaccinated individuals can still mount robust boosting in anti-N responses after SARS-CoV-2 infection, and that interpretation of anti-N measurement varied across studies due to the difference in the testing assay^6^.

Using longitudinal cohort data from the Household Influenza Vaccine Evaluation (HIVE) study and Community Vaccine Effectiveness (CoVE) study in Michigan, this study estimated post-infection changes in anti-N IgG concentrations and modeled antibody boosting and durability following first PCR-confirmed SARS-CoV-2 infections, overall and by age group.

## Methods

### Study Population and Lab Testing

Eligible participants were selected from the HIVE and the CoVE studies during periods of active SARS-CoV-2 surveillance. HIVE is an ongoing prospective household cohort that has followed participants since 2010, with SARS-CoV-2 surveillance conducted from January 2020 through July 2024. HIVE enrolled households living within 30 miles of the study clinic in Ann Arbor, Michigan, with at least three household members receiving primary care within the University of Michigan Health System and at least one child aged <18 years living in the household. CoVE enrolled and followed participants from September 2022 through July 2024. CoVE enrolled children and adults of all ages living in Michigan who received health care, with statewide recruitment through social media, the Michigan Medicine research registry, and outreach from health systems and community partners. Participants included in the present analysis experienced their eligible SARS-CoV-2 infections during these surveillance periods. Individual and household characteristics, self-report prior SARS-CoV-2 infection, as well as health history, were collected at enrollment. Detailed methods for the HIVE and CoVE have been previously published^7,8^.

Participants were instructed to self-collect a nasal swab upon onset of eligible illness, defined by ≥2 of the following symptoms: cough, fever, chills, nasal congestion, headache, body aches, or sore throat. A subset of participants self-collected nasal swabs weekly, regardless of symptoms, between September 2022-July 2024. Participants were asked to contribute blood specimens at the initial enrollment visit and at scheduled appointments twice annually between May and August and between October and January. Additional pre- and post-SARS-CoV-2 vaccination blood was collected approximately 14 days before and 7 and 28 days after SARS-CoV-2 vaccination^7^.

Nasal swabs were tested for SARS-CoV-2 using reverse transcription polymerase chain reaction (RT-qPCR). For specimens collected through October 2021, SARS-CoV-2 was identified using primers and probes developed by the Centers for Disease Control and Prevention (CDC); subsequent specimens were tested using the Thermo Fisher Scientific TaqPath COVID-19, Flu A, Flu B Combo Kit. Serum was tested for anti-N IgG antibody concentration with a multiplex electrochemiluminescence assay (Meso Scale Discovery, V-PLEX Respiratory Panel 4 IgG), lab procedure described elsewhere and in supplementary methods^7,9^. Absolute antibody quantity was measured in arbitrary units per milliliter (AU/mL) with a conversion factor of 0.00236 to Binding Antibody Unit per milliliter (BAU/mL). Per the assay insert, a concentration 5,000 AU/mL (corresponding to 11.8 BAU/mL) indicated infection positivity for anti-N IgG antibodies^7,9^.

This study was reviewed and approved by the University of Michigan Institutional Review Board and was conducted consistent with applicable federal law and CDC policy (See 45 C.F.R. part 46.114; 21 C.F.R. part 56.114).

### Eligibility Criteria

Participants were eligible for the overall study population if they had at least one blood sample collected <120 days after their first PCR-confirmed SARS-CoV-2 infection (with the infection date defined as the collection date of the first PCR-positive swab) and had no prior PCR-confirmed infection. A pre-infection serum sample was not required for inclusion in the overall study population. Positive PCR specimens collected within 30 days of each other were considered part of the same infection episode.

To evaluate antibody boosting following infection, participants were required to have both a pre-infection serum sample collected <180 days before infection and a post-infection serum sample collected <120 days after infection with anti-N IgG results available for both specimens. When multiple eligible pre-infection serum samples were available, the specimen collected closest to the infection date was selected for testing. The 180-day pre-infection window was chosen to capture the most recent baseline antibody measurement, whereas the 120-day post-infection window was selected to capture the early antibody response while minimizing overlap with subsequent infections. All post-infection serum samples collected within 120 days after infection were eligible for inclusion in the analysis.

To evaluate population-level anti-N IgG durability and the average change in antibody concentrations over time after infection, a pre-infection serum sample was not required. All eligible post-infection serum samples collected within 120 days after infection were initially considered to characterize population-level post-infection antibody kinetics. We subsequently restricted the durability analysis to post-infection serum samples collected from day 52 through day 120 after infection; this time period was chosen because day 52 was identified as the approximate population-level peak anti-N IgG response in our statistical analysis (Figure 1).

**Figure 1.**
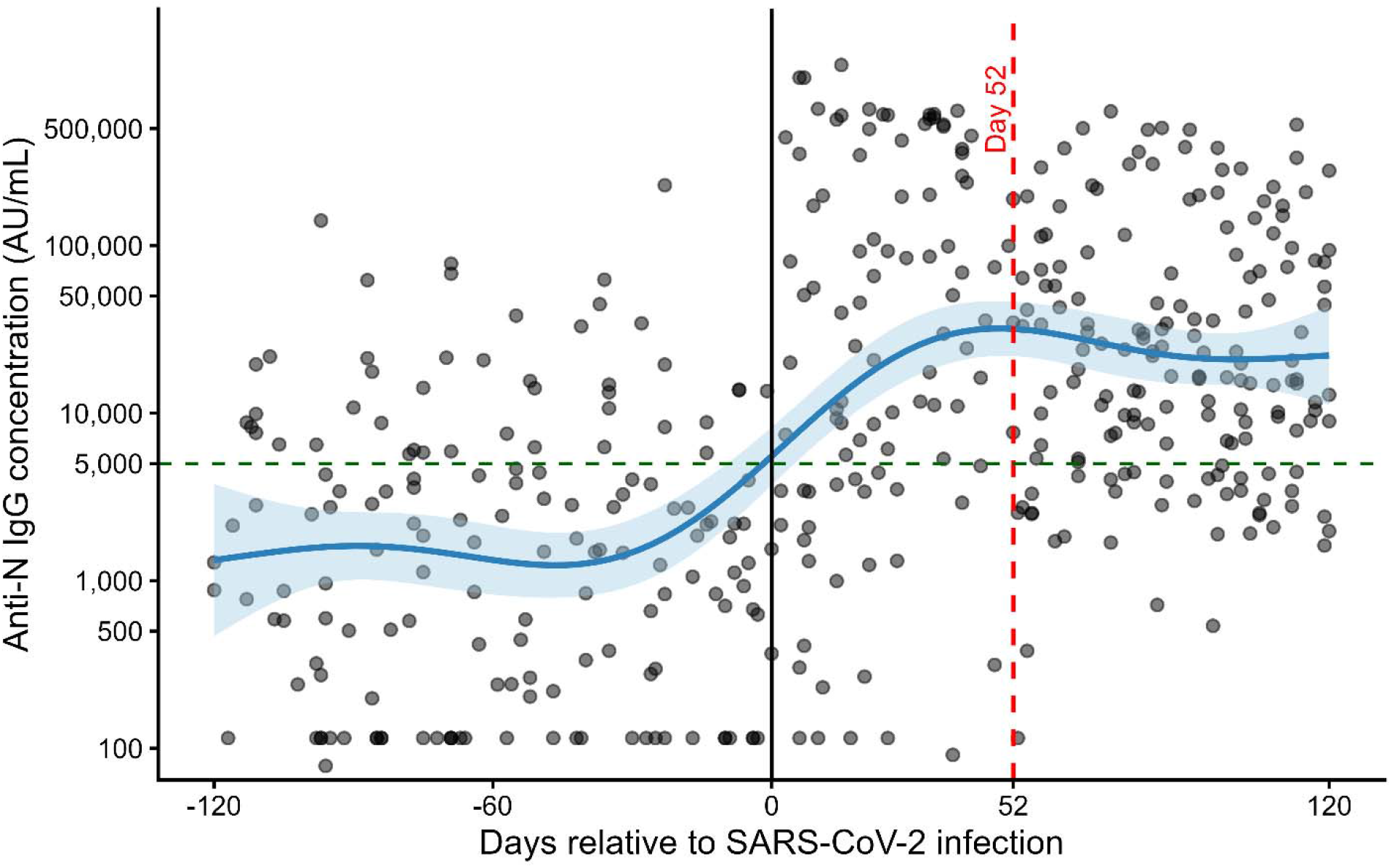
Anti-nucleocapsid IgG antibody concentrations before and after first SARS-CoV-2 infection (N=186). Anti-nucleocapsid (anti-N) IgG concentrations measured by electrochemiluminescence assay are shown relative to the date of participants’ first known PCR-confirmed SARS-CoV-2 infection (day 0), defined as the collection date of the first PCR-positive nasal swab. Each point represents a serum specimen, and the blue line represents a locally smoothed regression (LOESS) curve with the shaded area indicating the 95% confidence interval. The black vertical line denotes the date of infection (day 0), the red dashed line indicates day 52, corresponding to the observed peak anti-N IgG response and the start of the antibody waning analysis, and the green dashed horizontal line represents the seropositivity threshold (5,000 AU/mL).

### Statistical Analysis

Descriptive statistics were used to summarize demographic and COVID-19 vaccination information, stratified by age group (adults ≥18, minors <18). Anti-N IgG antibody concentration, which was used irrespective of anti-N seropositivity status, was log10-transformed. Geometric mean concentration (GMC) and, among participants with paired pre- and post-infection serum samples, geometric mean fold rise (GMFR) were calculated with standard 95% confidence intervals (CI) to assess changes in antibody concentrations following infection. Stratified analyses were performed by age, vaccine status, and pre-infection anti-N seropositivity status (i.e. from previously undetected infections).

To assess post-infection kinetics, a generalized additive model (GAM) was fit to log10-transformed anti-N IgG concentrations from all specimens collected <120 days post-infection, with time since infection modeled using a smooth function. The population-level peak was identified as the day with the highest predicted anti-N IgG concentration from the fitted curve. The estimated peak occurred at approximately day 52; therefore, day 52 was selected as the starting point based on the observed peak by that time (Figure 1), similar to a prior study^4^.

Antibody durability from day 52 through day 120 post-infection was then modeled using both unadjusted and adjusted linear mixed (LM) models. Household was included as a random effect to account for clustering of participants within households. The p-value for time tested the null hypothesis that the slope of log10 anti-N IgG concentration over days since infection was zero, corresponding to no change in antibody concentration over time. The adjusted model included age group (<18 vs. ≥18 years), presence of any high-risk health condition, and anti-N seropositivity prior to the first PCR-confirmed SARS-CoV-2 infection as fixed-effect covariates. Natural cubic spline (NCS) mixed models were also fit to characterize potential non-linear antibody waning trajectories, and likelihood ratio tests were used to compare NCS models with intercept-only mixed models. A 95% confidence interval was calculated using 100 nonparametric bootstrap replicates^10^. As a sensitivity analysis, we refit the models after randomly selecting one eligible serum specimen per participant to assess whether repeated observations from a subset of participants influenced the estimated population-level antibody trajectory. All analyses were conducted using R (version 4.5.1).

## Results

A total of 186 participants met the overall eligibility criteria, including 144 (77%) adults aged ≥18 years and 42 (23%) minors aged <18 years (Figure S1). Of these, 113 participants met the additional eligibility criteria for the antibody boosting analysis by having an eligible pre-infection serum specimen collected <180 days before infection and an eligible post-infection serum specimen collected <120 days after infection. A separate subset of 66 participants contributed serum specimens between days 52 and 120 and was included in the antibody durability analysis. The overall eligible study was predominantly White (80%), and non-Hispanic (94%), with a median age of 42 years (IQR: 33-60) (Table 1). About half (56%) reported at least one high-risk condition. Majority (81%) had received at least one dose of COVID-19 vaccine prior to their first PCR-confirmed SARS-CoV-2 infection.

**Table 1.** Characteristics of participants by age group, HIVE and CoVE cohorts, 2020-2024.

| Characteristic | Overall<br>N = 186 | <18 years<br>N = 42 | ≥18 years<br>N = 144 |
| --- | --- | --- | --- |
| **Age at infection, median (interquartile range [IQR]), years** | 42.0 (33.0, 60.0) | 12.0 (9.0, 15.0) | 47.0 (39.5, 63.0) |
| **Female** | 105 (56.5%) | 17 (40.5%) | 88 (61.1%) |
| **Race** |  |  |  |
| White | 149 (80.1%) | 36 (85.7%) | 113 (78.5%) |
| Asian | 6 (3.2%) | 1 (2.4%) | 5 (3.5%) |
| Biracial or multiracial | 4 (2.2%) | 1 (2.4%) | 3 (2.1%) |
| Black or African American | 1 (0.5%) | 1 (2.4%) | 0 (0.0%) |
| Middle Eastern or North African | 2 (1.1%) | 0 (0.0%) | 2 (1.4%) |
| Missing or unknown | 24 (12.9%) | 3 (7.1%) | 21 (14.6%) |
| **Ethnicity** |  |  |  |
| Hispanic or Latino | 12 (6.5%) | 5 (11.9%) | 7 (4.9%) |
| **Year of enrollment** |  |  |  |
| 2020 or earlier | 55 (30.9%) | 16 (39.0%) | 39 (28.5%) |
| 2021 | 13 (7.3%) | 5 (12.2%) | 8 (5.8%) |
| 2022 | 14 (7.9%) | 3 (7.3%) | 11 (8.0%) |
| 2023 | 96 (53.9%) | 17 (41.5%) | 79 (57.7%) |
| Missing | 8 | 1 | 7 |
| **Any high-risk health condition<sup>1</sup>** | 104 (55.9%) | 10 (23.8%) | 94 (65.3%) |
| **Vaccinated before infection<sup>2</sup>** | 151 (81.2%) | 31 (73.8%) | 120 (83.3%) |
| **Vaccinated within 6 months before infection** | 81 (43.5%) | 14 (33.3%) | 67 (46.5%) |
| **# vaccine doses before infection, median (IQR)<sup>3</sup>** | 4.0 (3.0, 5.0) | 3.0 (0.0, 4.0) | 4.0 (3.0, 6.0) |
| **Anti-N IgG serostatus before infection<sup>4</sup>** |  |  |  |
| Seronegative | 82 (44.1%) | 13 (31.0%) | 69 (47.9%) |
| Seropositive | 31 (16.7%) | 8 (19.0%) | 23 (16.0%) |
| Unknown | 73 (39.2%) | 21 (50.0%) | 52 (36.1%) |
| **Self-report SARS-CoV-2 infection prior to the CoVE cohort enrollment, n/N (%)**<sup>5</sup> | 30/157 (19.1%) | 8/35 (22.9%) | 22/122 (18.0%) |
| **Time since self-reported pre-enrollment SARS-CoV-2 infection date to pre first PCR-confirmed infection serum collection date after enrollment in the CoVE study, median (IQR), days** | 438 (234-543) | 550 (496-610) | 464 (336-576) |

<sup>1</sup> High-risk health conditions included chronic obstructive pulmonary disease, sleep apnea, cardiac disease, heart failure, hypertension, diabetes, malignancy, arthritis, stroke, deep vein thrombosis/pulmonary embolism, immunosuppressive conditions, depression, chronic kidney disease, liver disease, blood disorders, neurologic disorders, endocrine disorders, and anxiety.
<sup>2</sup> Participants had documented COVID-19 vaccine doses received before their first known PCR-confirmed SARS-CoV-2 infection.
<sup>3</sup> Total number of documented COVID-19 vaccine doses received before the participant's first known PCR-confirmed SARS-CoV-2 infection.
<sup>4</sup> Anti-N IgG serostatus was defined using the electrochemiluminescence assay; per the manufacturer's instructions, participants with anti-nucleocapsid IgG concentrations ≥5,000 AU/mL were classified as seropositive. Unknown indicates those participants who did not have serological sample collection within <180 days of their first PCR-confirmed infection.
<sup>5</sup> participants enrolled in the CoVE cohort (started September 2022) provided self-reported SARS-CoV-2 infection prior to enrollment. n= number of participants reported infection before enrollment, N=total number of participants have non-missing self-report prior infection status

Among 113 participants included in the antibody boosting analysis, 28% of participants were anti-N seropositive before infection and 80% of participants were anti-N seropositive after infection (Table S1-S2). Pre-infection samples were collected a median of 69 days (IQR: 30–106) before infection, and post-infection samples were collected a median of 55 days (IQR: 21–84) after infection. The pre-infection geometric mean concentration (GMC) of SARS-CoV-2 anti-N IgG antibodies was 1,098.0 AU/mL (95% CI: 745.0, 1,618.3), increasing to a post-infection GMC of 25,417.5 AU/mL (95% CI: 16,766.0, 38,533.4) (Figure 2, Table S1-S2). The overall geometric mean fold rise (GMFR) following infection was 23.15 (95% CI: 15.61, 34.33). Overall, 84.1% experienced at least a two-fold increase in anti-N IgG concentrations, and 80.5% experienced at least a four-fold increase.

**Figure 2.**
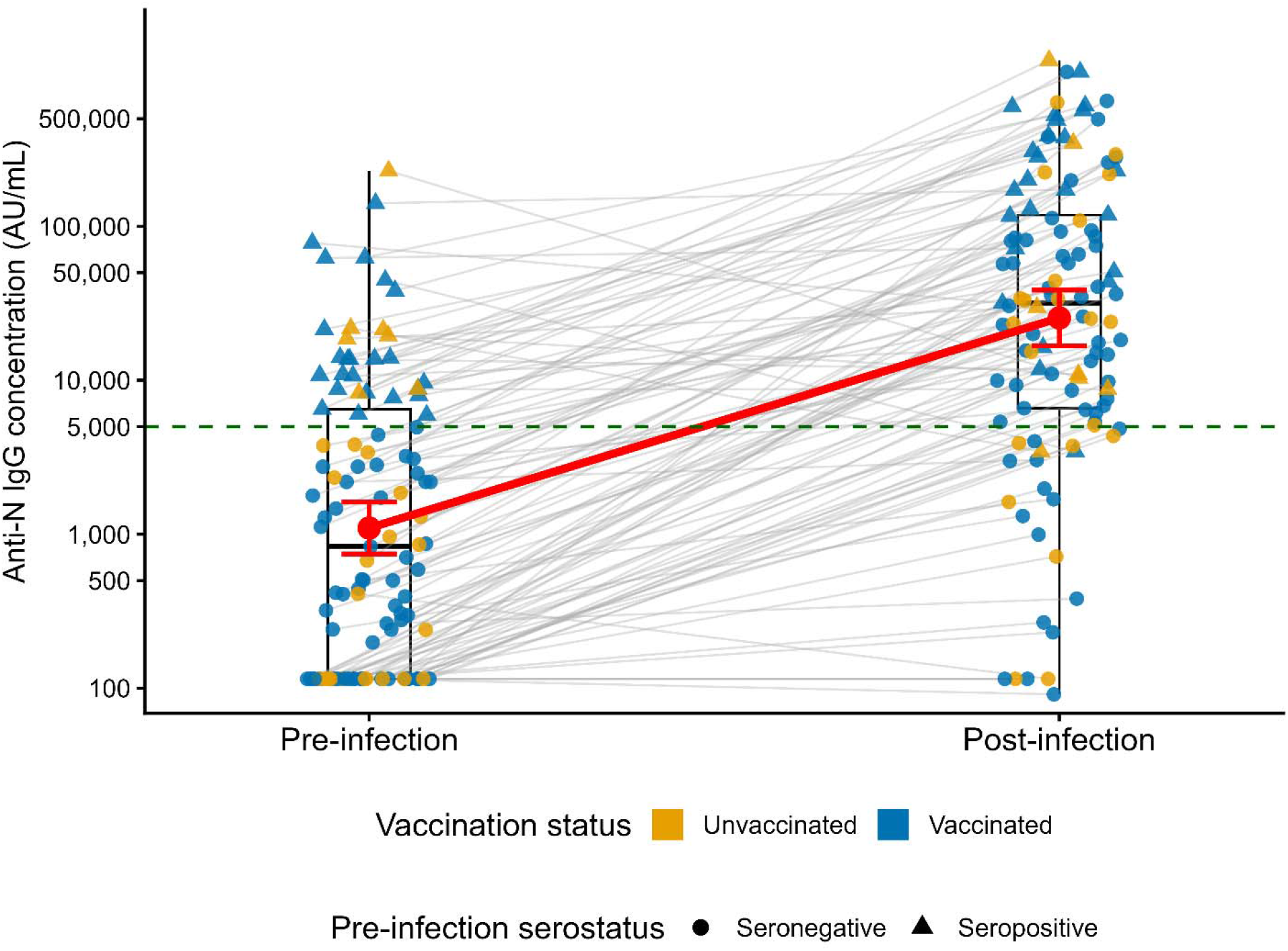
Anti-nucleocapsid IgG antibody responses before and after first SARS-CoV-2 infection (N=113). Anti-nucleocapsid (anti-N) IgG concentrations measured by electrochemiluminescence (ECL) assay among participants with paired pre- and post-infection serum specimens. Each gray line connects paired serum samples from an individual participant. Points are colored by COVID-19 vaccination status before infection (blue, vaccinated; gold, unvaccinated) and shaped by pre-infection anti-N IgG serostatus (circles, seronegative; triangles, seropositive). Boxplots show the median (center line), interquartile range (box), and values extending to 1.5 times the interquartile range (whiskers). The red line and points represent the geometric mean anti-N IgG concentrations before and after infection, with error bars indicating 95% confidence intervals. The green dashed horizontal line denotes the anti-N IgG seropositivity threshold (5,000 AU/mL) as determined by the manufacturer.

We next assessed pre- and post-infection anti-N IgG concentrations and fold increases among these 113 participants, stratified by age group, prior vaccination status and pre-infection anti-N serostatus. We did not observe differences in antibody boosting when stratified by age group (adults vs minors). Among 92 adults (≥18 years), the pre-infection GMC was 1,084.8 AU/mL (95% CI: 715.1, 1,645.5) and the post-infection GMC was 27,863.8 AU/mL (95% CI: 17,325.6, 44,812.1), corresponding to a GMFR of 25.69 (95% CI: 16.64, 39.65). Among 21 minors (<18 years), the pre-infection GMC was 1,157.9 AU/mL (95% CI: 385.8, 3,474.9) and the post-infection GMC was 16,994.1 AU/mL (95% CI: 7,041.8, 41,011.8), corresponding to a GMFR of 14.68 (95% CI: 5.43, 39.69). Stratified by vaccination status, we did not observe a difference in antibody boosting among vaccinated (GMFR: 26.19 [95% CI: 17.30, 39.64]) and unvaccinated participants (GMFR: 15.92 [95% CI: 5.76, 44.00]) (Table S3). In contrast, participants who were seronegative before infection demonstrated substantially greater antibody boosting (GMFR: 38.73 [95% CI: 25.72, 58.33]) than those who were seropositive before infection (GMFR: 5.93 [95% CI: 2.68, 13.12]), although post-infection seropositivity exceeded 70% in both groups (Table S4).

To estimate antibody durability at the population level, we analyzed 84 post-infection serum specimens contributed by 66 participants between days 52 and 120 after infection (Table S5). Because most participants contributed a single post-infection specimen and only a small number contributed repeated measurements, the estimated antibody waning primarily reflects the population-average trajectory rather than individual longitudinal antibody changes. In the unadjusted LM model, anti-N IgG concentrations did not show a significant monthly decline (11.3% per month, 95% CI: -8.4% to 27.5%; p = 0.228) (Figure 3A). Results were similar after adjustment for age group, high-risk health conditions, and pre-infection seropositivity (Table S6).

**Figure 3.**
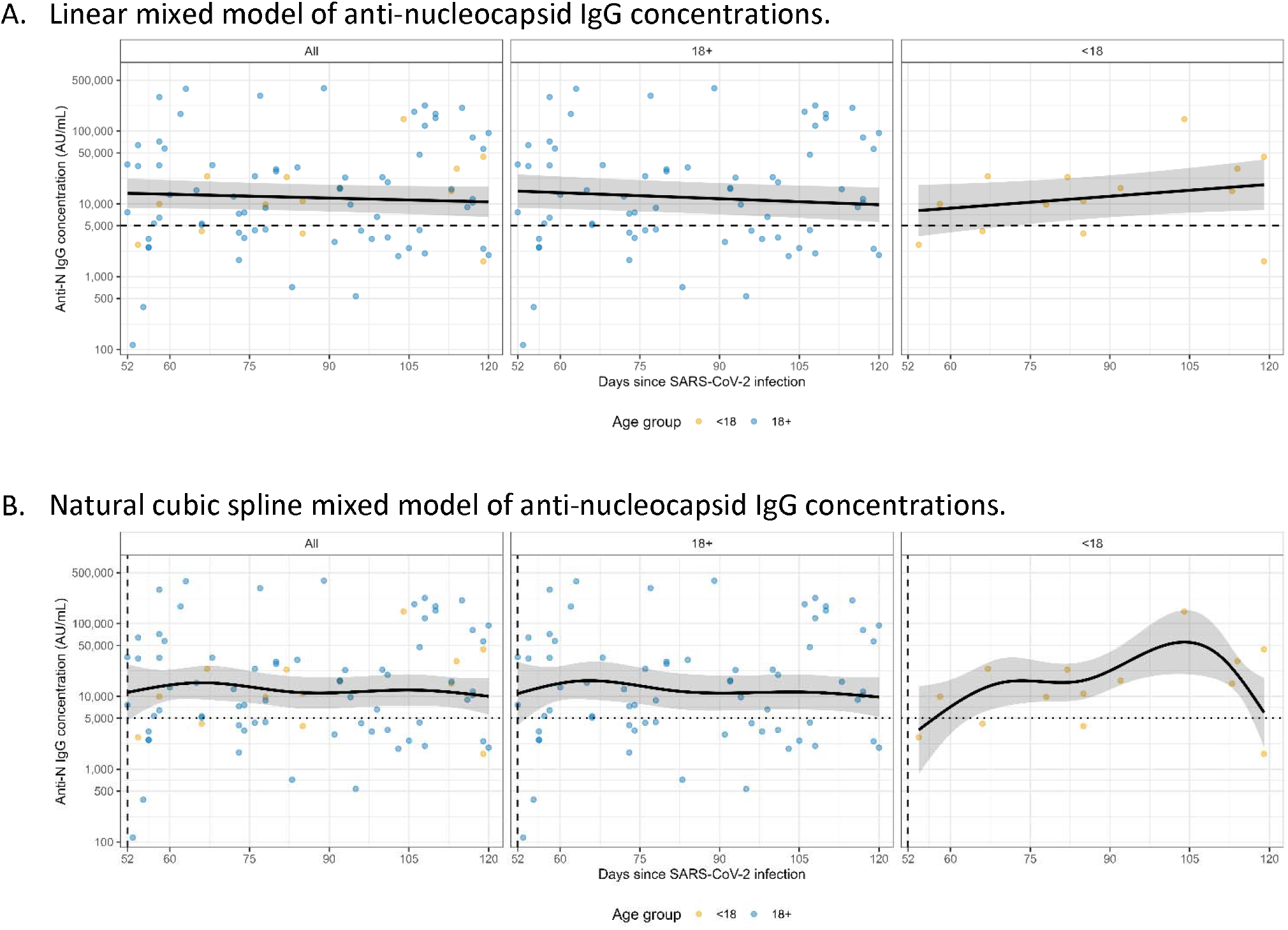
Trajectories of anti-N IgG concentration post SARS-CoV-2 PCR-confirmed infection (N=66). Anti-nucleocapsid (anti-N) IgG concentrations among participants with post-infection serum specimens collected between 52 and 120 days after first known PCR-confirmed SARS-CoV-2 infection, with the infection date (day 0) defined as the collection date of the first PCR-positive nasal swab. Results are shown for all participants, adults (≥18 years), and minors (<18 years). Points represent individual serum specimens. Solid lines represent the estimated population-level trajectories, and shaded areas indicate 95% confidence intervals. Panel A shows estimates from an unadjusted linear mixed model, whereas Panel B shows estimates from an unadjusted natural cubic spline mixed model to assess potential non-linear changes in antibody concentrations over time. The horizontal dashed line indicates the anti-N IgG seropositivity threshold (5,000 AU/mL).

In age-stratified models, adults demonstrated durable population-level anti-N IgG antibody responses, with an estimated 17.3% monthly decline in antibody concentrations (95% CI: -1.5% to 32.6%; p = 0.067, Figure 3A, Table S6). Among minors, model estimates were highly imprecise, with wide confidence intervals encompassing both increases and decreases in antibody concentrations over time, likely reflecting the limited sample size (Figure 3A, Table S6). We also fit NCS models to explore non-linear antibody trajectories and showed similar trends as the linear model (Figure 3B). Sensitivity analyses similarly showed no statistically significant decline in anti-N IgG concentrations overall or among adults (Table S7).

## Discussion

In this study, we observed robust boosting of anti-N IgG antibodies following first PCR-confirmed SARS-CoV-2 infection, including those with previous vaccination. More than 80% of participants experienced at least a four-fold increase in anti-N IgG concentrations following infection, and approximately 80% had post-infection titers above the manufacturer’s threshold for seropositivity. We then modeled the durability of anti-nucleocapsid IgG antibodies following SARS-CoV-2 infection using both linear mixed models and natural cubic spline mixed models. These approaches allowed us to evaluate both overall trends and potential non-linear antibody trajectories over time. Population-level analyses demonstrated sustained anti-N IgG concentrations between 52 and 120 days after infection, with no evidence of substantial decline.

Although anti-N antibodies are useful markers of prior SARS-CoV-2 infection, their relationship with protection against subsequent infection is less well established than that of spike-specific binding and neutralizing antibodies. Therefore, the magnitude and durability of anti-N responses observed in this study should primarily inform interpretation of serologic evidence of prior infection rather than be interpreted as a direct measure of protective immunity. Consistent with prior studies, our results suggest that SARS-CoV-2 infection induces substantial boosting of anti-N antibody responses, including among individuals vaccinated before infection^3,11–16^. More than 80% of participants experienced at least a four-fold increase in anti-N IgG concentrations following infection, supporting the utility of anti-N antibodies as markers of prior infection in serologic studies. Although antibody boosting was observed regardless of vaccination status, participants who were anti-N seronegative before infection exhibited substantially greater fold rises than those who were seropositive before infection, likely reflecting higher baseline anti-N IgG concentrations among previously seropositive individuals and the smaller relative increase achievable after reinfection. In addition, more than one-quarter of participants were anti-N seropositive before their first PCR-confirmed SARS-CoV-2 infection, suggesting prior undocumented mild or asymptomatic infection. This finding highlights the limitations of relying solely on molecular surveillance to define first infection and indicates the value of integrating serologic and molecular data to more completely capture SARS-CoV-2 exposure. Furthermore, baseline anti-N seropositivity was associated with smaller post-infection antibody boosts, indicating that pre-existing anti-N antibody levels should be considered when interpreting longitudinal changes in antibody concentrations in serosurveillance studies.

Strength of this study include the use of repeated serologic measurements in a well-characterized prospective cohort and the application of flexible spline-based modeling to capture antibody dynamics over time. In addition, although the study protocol did not specifically include scheduled post-infection blood draws, the longitudinal cohort design with repeated routine specimen collection allowed us to capture a substantial number of paired pre- and post-infection samples across a wide range of post-infection time points. Several limitations should be considered when interpreting our findings. First, participants were enrolled and followed during 2020–2024, and their baseline immunity may not reflect current population immunity after additional years of SARS-CoV-2 circulation and repeated infections. Individuals currently experiencing infection may be more likely to have pre-existing anti-N antibodies, similar to the participants in our study who were seropositive before their first known PCR-confirmed infection. Because pre-infection seropositive participants demonstrated smaller antibody increases following infection, our findings may underestimate the magnitude of anti-N antibody boosting expected following their first SARS-CoV-2 infection among populations who were infection naïve. The lack of racial, ethnic, and geographic diversity in the sample, which was predominantly White, non-Hispanic, and female. Our follow-up was also limited to 120 days after infection, preventing assessment of longer-term antibody persistence or subsequent declines^15,17^. Most participants in the durability analysis contributed only one post-infection specimen, thus the estimated waning primarily represents a population-average trajectory rather than within-person longitudinal antibody decline. The number of pediatric participants was also small, further limiting our ability to evaluate age-specific antibody responses and durability. Another limitation is that the number of pediatric participants was small, limiting our ability to directly compare antibody responses and durability between children and adults. Consequently, our age-stratified analyses should be interpreted with caution, and the overall findings are largely driven by the adult population. Moreover, our modeling did not account for vaccine type or timing, which may have resulted in residual confounding if vaccination history influenced baseline or post-infection anti-N antibody responses.

In conclusion, SARS-CoV-2 infection elicited robust anti-N IgG antibody responses, with most participants demonstrating substantial increases in antibody concentrations and seroconversion following infection. Population-level analyses indicated that anti-N IgG concentrations remained durable through at least 120 days after infection, supporting the use of anti-N antibodies as markers of recent SARS-CoV-2 infection in seroepidemiologic studies, including in highly vaccinated populations. However, the magnitude of antibody boosting varied according to pre-infection serostatus, suggesting that baseline antibody levels should be considered when interpreting serologic boosting as evidence of infection. These findings provide evidence to inform the design and interpretation of longitudinal SARS-CoV-2 serosurveillance studies in populations with heterogeneous infection and vaccination histories.

## Ethical statement

The University of Michigan Institutional Review Board gave ethical approval for this work. Informed consent was obtained at initial enrollment. Parents and legal guardians provided written informed consent for both themselves and their dependents. Children ages 7-9 provided consent verbally, while those 10-13 signed a separate, age-appropriate assent document. Adolescents ages 14-17 read and signed the full informed consent document.

## Finding statement

This study was supported by funding from the National Center for Immunization and Respiratory Diseases, US Centers for Disease Control and Prevention (75D30122C13149), and the National Institute of Allergy and Infectious Diseases and the National Institutes of Health (75N93021C00015).

## Disclaimer

The Centers for Disease Control and Prevention provided input into the study design and data collection for the overall HIVE and CoVE cohort. The funders did not have any role in data analysis, interpretation, or writing of the manuscript. The Centers for Disease Control and Prevention reviewed and approved the draft manuscript before submission. The findings and conclusions in this report are those of the author(s) and do not necessarily represent the official position of the Centers for Disease Control and Prevention. The National Institutes of Health received a final copy of the manuscript prior to submission.

## Data availability

The data used in this study can be made available upon request. Due to Institutional Review Board (IRB) regulations, data access is controlled. Per the guidelines of the Centers for Excellence in Influenza Research and Response (CEIRR) Network, individuals seeking access must complete a data and specimen collaboration form. Requests received will be reviewed by the study investigators. For enquiries, please contact. Requests will generally receive an initial response within 3–5 business days.

## Supplementary Methods

### SARS-CoV-2 anti-nucleocapsid IgG measurement

Serum SARS-CoV-2 nucleocapsid (N)-specific IgG antibodies were measured using the V-PLEX COVID-19 Serology Panel 4 multiplex electrochemiluminescence (ECL) assay (Meso Scale Discovery [MSD], Rockville, MD, USA), according to the manufacturer’s protocol and established laboratory procedures. Serum specimens were diluted in MSD Diluent 100 and tested on multi-spot plates containing immobilized SARS-CoV-2 antigens. Antigen-specific IgG was detected using a sulfo-tag-conjugated anti-human IgG antibody, and ECL signals were measured using an MSD instrument. A serum-based MSD reference standard and assay controls were included on each plate. A seven-point calibration series with four-fold serial dilutions and a zero-calibrator blank was used to generate plate-specific calibration curves. Anti-N IgG concentrations were determined by back-fitting ECL signals to the calibration curve and correcting for specimen dilution and were reported in arbitrary units per milliliter (AU/mL). Anti-N IgG concentrations ≥5,000 AU/mL were classified as seropositive based on the manufacturer-established cutoff. The cutoff was established using pre-2019 specimens and specimens from individuals with PCR-confirmed COVID-19; at ≥15 days following PCR-confirmed infection, the manufacturer reported 93.8% sensitivity and 100% specificity. Anti-N IgG concentrations were log10-transformed for statistical analyses.

**Figure S1.**
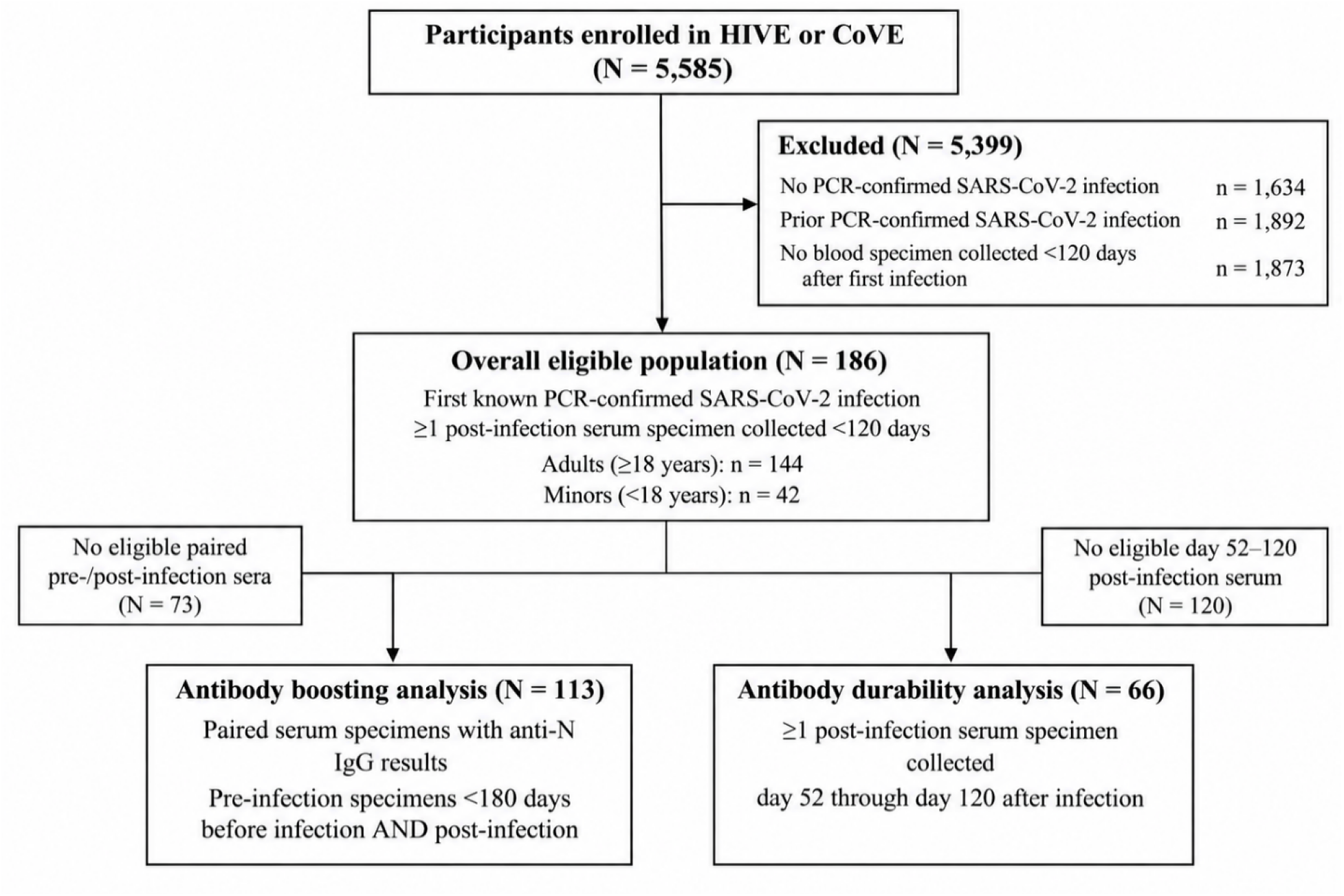
Participant flow and analytic populations.

**Table S1.** Characteristics of participants with paired pre- and post-infection serum specimens.

| Characteristic | Overall<br>N=113 | <18 years<br>N = 21 | ≥18 years<br>N = 92 |
| --- | --- | --- | --- |
| **Age at infection, mean (SD), years** | 45.4 (21.3) | 10.8 (5.0) | 53.3 (14.5) |
| **Sex** |  |  |  |
| Female | 59 (52.2%) | 9 (42.9%) | 50 (54.3%) |
| Male | 36 (31.9%) | 11 (52.4%) | 25 (27.2%) |
| Missing | 18 | 1 | 17 |
| **Race** |  |  |  |
| Asian | 2 (1.8%) | 1 (4.8%) | 1 (1.1%) |
| Biracial or multiracial | 1 (0.9%) | 1 (4.8%) | 0 (0.0%) |
| Black or African American | 0 (0.0%) | 0 (0.0%) | 0 (0.0%) |
| Middle Eastern or North African | 1 (0.9%) | 0 (0.0%) | 1 (1.1%) |
| Missing or unknown | 18 (15.9%) | 1 (4.8%) | 17 (18.5%) |
| White | 91 (80.5%) | 18 (85.7%) | 73 (79.3%) |
| **Ethnicity** |  |  |  |
| Hispanic or Latino | 5 (4.4%) | 2 (9.5%) | 3 (3.3%) |
| **Any high-risk health condition<sup>1</sup>** | 66 (58.4%) | 6 (28.6%) | 60 (65.2%) |
| **Vaccinated before infection<sup>2</sup>** | 85 (75.2%) | 13 (61.9%) | 72 (78.3%) |
| **# vaccine doses before infection, median (IQR)<sup>3</sup>** | 3.0 (2.0, 6.0) | 2.0 (0.0, 3.0) | 4.0 (2.5, 6.0) |
| **Anti-N IgG serostatus before infection<sup>4</sup>** |  |  |  |
| Seronegative | 82 (72.6%) | 13 (61.9%) | 69 (75.0%) |
| Seropositive | 31 (28.4%) | 8 (38.1%) | 23 (25.0%) |

<sup>1</sup> High-risk health conditions included chronic obstructive pulmonary disease, sleep apnea, cardiac disease, heart failure, hypertension, diabetes, malignancy, arthritis, stroke, deep vein thrombosis/pulmonary embolism, immunosuppressive conditions, depression, chronic kidney disease, liver disease, blood disorders, neurologic disorders, endocrine disorders, and anxiety.
<sup>2</sup> Participants had documented COVID-19 vaccine doses received before their first PCR-confirmed SARS-CoV-2 infection.
<sup>3</sup> Total number of documented COVID-19 vaccine doses received before the participant's first PCR-confirmed SARS-CoV-2 infection.
<sup>4</sup> Anti-N IgG serostatus was defined using the electrochemiluminescence assay; participants with anti-nucleocapsid IgG concentrations ≥5,000 AU/mL were classified as seropositive.

**Table S2:**
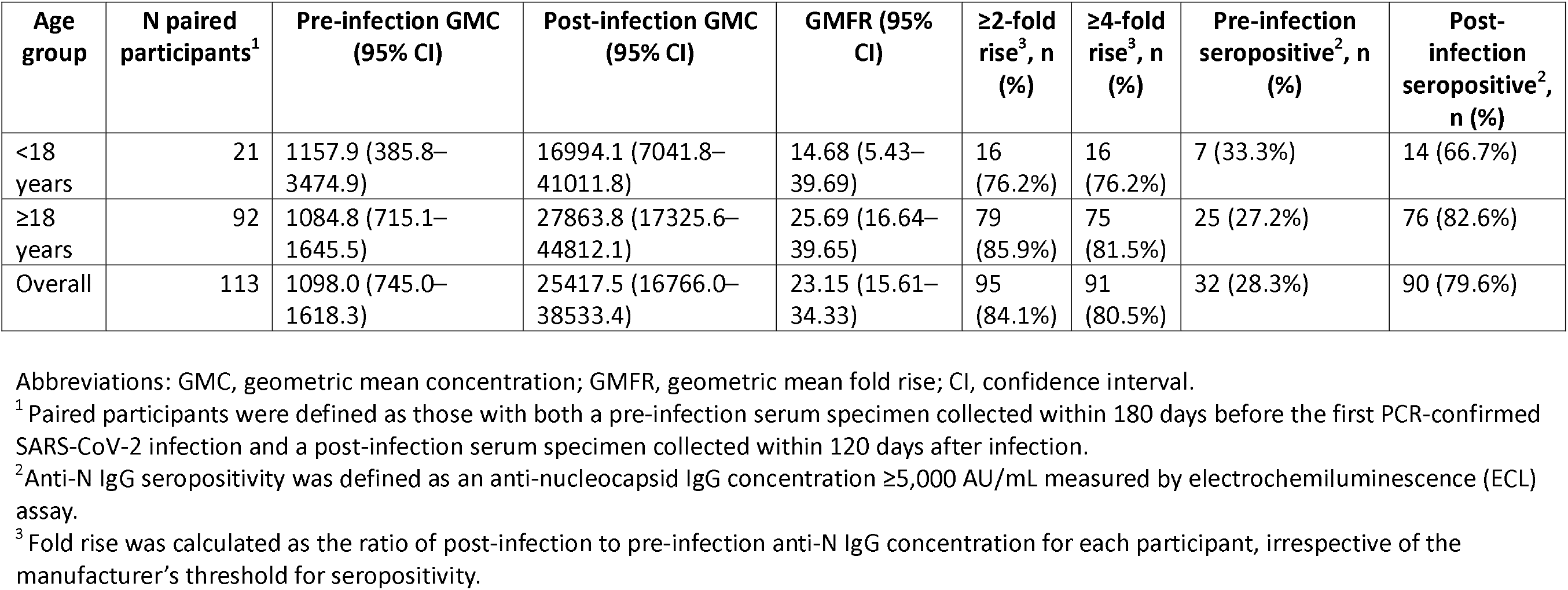
Pre- and post-infection geometric mean anti-nucleocapsid IgG concentrations and antibody responses among participants with paired serum specimens, overall and by age group.

**Table S3.** Pre- and post-infection geometric mean anti-nucleocapsid IgG concentrations and antibody responses among participants with paired serum specimens, stratified by COVID-19 vaccination status before first SARS-CoV-2 infection.

| Vaccination status<sup>1</sup> | Age group | N paired participants<sup>2</sup> | Pre-infection GMC (95% CI) | Post-infection GMC (95% CI) | GMFR (95% CI) | ≥2-fold rise<sup>4</sup>, n (%) | ≥4-fold rise<sup>4</sup>, n (%) | Pre-infection seropositive<sup>3</sup>, n (%) | Post-infection seropositive<sup>3</sup>, n (%) |
| --- | --- | --- | --- | --- | --- | --- | --- | --- | --- |
| Vaccinated | <18 years | 13 | 791.7<br>(187.5–3341.8) | 20178.6<br>(5629.7–72326.3) | 25.49<br>(8.34–77.89) | 11<br>(84.6%) | 11<br>(84.6%) | 4 (30.8%) | 9 (69.2%) |
|  | ≥18 years | 72 | 1161.3<br>(725.4–1859.0) | 30560.4<br>(18097.2–51606.9) | 26.32<br>(16.65–41.58) | 63<br>(87.5%) | 59<br>(81.9%) | 20 (27.8%) | 61 (84.7%) |
|  | Overall | 85 | 1095.2<br>(703.2–1705.7) | 28680.6<br>(17825.1–46147.3) | 26.19<br>(17.30–39.64) | 74<br>(87.1%) | 70<br>(82.4%) | 24 (28.2%) | 70 (82.4%) |
| Unvaccinated | <18 years | 8 | 2147.9<br>(267.3–17257.1) | 12855.3<br>(3089.4–53491.7) | 5.99 (0.73–48.83) | 5<br>(62.5%) | 5<br>(62.5%) | 4 (50.0%) | 5 (62.5%) |
|  | ≥18 years | 20 | 848.8<br>(320.6–2247.5) | 19980.9<br>(6067.1–65803.3) | 23.54<br>(6.90–80.36) | 16<br>(80.0%) | 16<br>(80.0%) | 3 (15.0%) | 15 (75.0%) |
|  | Overall | 28 | 1106.7<br>(471.6–2596.9) | 17615.4<br>(7178.6–43225.6) | 15.92<br>(5.76–44.00) | 21<br>(75.0%) | 21<br>(75.0%) | 7 (25.0%) | 20 (71.4%) |

Abbreviations: GMC, geometric mean concentration; GMFR, geometric mean fold rise; CI, confidence interval.
<sup>1</sup> Vaccination status was defined according to receipt of at least one COVID-19 vaccine dose before the participant's first PCR-confirmed SARS-CoV-2 infection.
<sup>2</sup> Paired participants were defined as those with both a pre-infection serum specimen collected within 180 days before the first PCR-confirmed SARS-CoV-2 infection and a post-infection serum specimen collected within 120 days after infection.
<sup>3</sup> Anti-N IgG seropositivity was defined as an anti-nucleocapsid IgG concentration ≥5,000 AU/mL measured by electrochemiluminescence (ECL) assay.
<sup>4</sup> Fold rise was calculated as the ratio of post-infection to pre-infection anti-N IgG concentration for each participant.

**Table S4.** Pre- and post-infection geometric mean anti-nucleocapsid IgG concentrations and antibody responses among participants with paired serum specimens, stratified by pre-infection anti-N IgG seropositivity status.

| Pre-infection anti-N IgG seropositivity status<sup>1</sup> | Age group | N paired participants<sup>2</sup> | Pre-infection GMC (95% CI) | Post-infection GMC (95% CI) | GMFR (95% CI) | ≥2-fold rise<sup>4</sup>, n (%) | ≥4-fold rise<sup>4</sup>, n (%) | Pre-infection seropositive<sup>3</sup>, n (%) | Post-infection seropositive<sup>3</sup>, n (%) |
| --- | --- | --- | --- | --- | --- | --- | --- | --- | --- |
| Seropositive | <18 years | 8 | 17878.9<br>(10805.5–29582.7) | 57945.4<br>(9486.0–353962.6) | 3.24<br>(0.60–17.43) | 4<br>(50.0%) | 4<br>(50.0%) | 8 (100.0%) | 7 (87.5%) |
|  | ≥18 years | 23 | 17131.7<br>(10907.2–26908.2) | 125399.9<br>(65410.5–240406.9) | 7.32<br>(2.81–19.10) | 16<br>(69.6%) | 16<br>(69.6%) | 23 (100.0%) | 22 (95.7%) |
|  | Overall | 31 | 17321.5<br>(12267.9–24456.8) | 102748.1<br>(55181.3–191318.0) | 5.93<br>(2.68–13.12) | 20<br>(64.5%) | 20<br>(64.5%) | 31 (100.0%) | 29 (93.5%) |
| Seronegative | <18 years | 13 | 214.9<br>(104.2–443.2) | 7988.6<br>(3481.2–18332.0) | 37.18<br>(12.74–108.50) | 12<br>(92.3%) | 12<br>(92.3%) | 0 (0.0%) | 7 (53.8%) |
|  | ≥18 years | 69 | 432.4<br>(318.2–587.6) | 16876.8<br>(9711.8–29327.6) | 39.03<br>(24.77–61.50) | 63<br>(91.3%) | 59<br>(85.5%) | 0 (0.0%) | 54 (78.3%) |
|  | Overall | 82 | 387.0<br>(291.9–513.1) | 14989.7<br>(9263.6–24255.3) | 38.73<br>(25.72–58.33) | 75<br>(91.5%) | 71<br>(86.6%) | 0 (0.0%) | 61 (74.4%) |

Abbreviations: GMC, geometric mean concentration; GMFR, geometric mean fold rise; CI, confidence interval.
<sup>1</sup> Pre-infection anti-N IgG serostatus was determined using the pre-infection serum specimen. Participants with anti-nucleocapsid IgG concentrations ≥5,000 AU/mL measured by electrochemiluminescence (ECL) assay were classified as seropositive; those with concentrations <5,000 AU/mL were classified as seronegative.
<sup>2</sup> Paired participants were defined as those with both a pre-infection serum specimen collected within 180 days before the first PCR-confirmed SARS-CoV-2 infection and a post-infection serum specimen collected within 120 days after infection.
<sup>3</sup> Anti-N IgG seropositivity was defined as an anti-nucleocapsid IgG concentration ≥5,000 AU/mL measured by electrochemiluminescence (ECL) assay.
<sup>4</sup> Fold rise was calculated as the ratio of post-infection to pre-infection anti-N IgG concentration for each participant.

**Table S5.** Characteristics of participants included in the antibody waning analysis.

| Characteristic | Overall<br>N=66 | <18 years<br>N = 12<sup>1</sup> | ≥18 years<br>N = 54<sup>1</sup> |
| --- | --- | --- | --- |
| **Age at infection, mean (SD), years** | 44.4 (20.1) | 12.1 (3.8) | 51.6 (14.3) |
| **Sex** |  |  |  |
| Female | 34 (51.5%) | 7 (58.3%) | 27 (50.0%) |
| Male | 20 (30.3%) | 5 (41.7%) | 15 (27.8%) |
| Missing | 12 | 0 | 12 |
| **Race** |  |  |  |
| Asian | 2 (3.0%) | 0 (0.0%) | 2 (3.7%) |
| Biracial or multiracial | 2 (3.0%) | 1 (8.3%) | 1 (1.9%) |
| Black or African American | 0 (0.0%) | 0 (0.0%) | 0 (0.0%) |
| Middle Eastern or North African | 0 (0.0%) | 0 (0.0%) | 0 (0.0%) |
| Missing or unknown | 12 (18.2%) | 0 (0.0%) | 12 (22.2%) |
| White | 50 (75.8%) | 11 (91.7%) | 39 (72.2%) |
| **Ethnicity** |  |  |  |
| Hispanic or Latino | 4 (6.1%) | 2 (16.7%) | 2 (3.7%) |
| **Any high-risk health condition<sup>1</sup>** | 42 (63.6%) | 7 (58.3%) | 35 (64.8%) |
| **Vaccinated before infection<sup>2</sup>** | 47 (71.2%) | 7 (58.3%) | 40 (74.1%) |
| **# vaccine doses before infection, median (IQR)<sup>3</sup>** | 3.0 (0.0, 5.0) | 2.0 (0.0, 3.0) | 3.0 (0.0, 6.0) |
| **Anti-N IgG serostatus before infection<sup>4</sup>** |  |  |  |
| Seronegative | 47 (81.0%) | 9 (81.8%) | 38 (80.9%) |
| Seropositive | 11 (19.0%) | 2 (18.2%) | 9 (19.1%) |
| Unknown | 8 | 1 | 7 |

<sup>1</sup> High-risk health conditions included chronic obstructive pulmonary disease, sleep apnea, cardiac disease, heart failure, hypertension, diabetes, malignancy, arthritis, stroke, deep vein thrombosis/pulmonary embolism, immunosuppressive conditions, depression, chronic kidney disease, liver disease, blood disorders, neurologic disorders, endocrine disorders, and anxiety.
<sup>2</sup> Participants had documented COVID-19 vaccine doses received before their first PCR-confirmed SARS-CoV-2 infection.
<sup>3</sup> Total number of documented COVID-19 vaccine doses received before the participant's first PCR-confirmed SARS-CoV-2 infection.
<sup>4</sup> Anti-N IgG serostatus was defined using the electrochemiluminescence assay; participants with anti-nucleocapsid IgG concentrations ≥5,000 AU/mL were classified as seropositive.

**Table S6.** Linear Mixed Model Starting at 52 Days Following SARS-CoV-2 Infection.

| Population | Model | Slope/day (95% CI) | Monthly decline % (95% CI) | P-value |
| --- | --- | --- | --- | --- |
| All | Unadjusted | -0.0017 (-0.0047, 0.0012) | 11.3 (-8.4, 27.5) | 0.228 |
| All | Adjusted | -0.0017 (-0.0046, 0.0012) | 11.3 (-8.4, 27.3) | 0.229 |
| <18 | Unadjusted | 0.0055 (-0.0043, 0.0152) | -45.8 (-185.6, 25.6) | 0.175 |
| <18 | Adjusted | 0.0060 (-0.0048, 0.0168) | -50.9 (-218.5, 28.5) | 0.170 |
| 18+ | Unadjusted | -0.0027 (-0.0057, 0.0002) | 17.3 (-1.5, 32.6) | 0.067 |
| 18+ | Adjusted | -0.0029 (-0.0058, 0.0001) | 18.0 (-0.4, 33.0) | 0.055 |

**Table S7:** Sensitivity Analysis of Anti-Nucleocapsid IgG Antibody Trajectories Among Participants Contributing a Single Serum Specimen Between 52 and 120 Days Following SARS-CoV-2 Infection.

| Population | Model | Slope/day (95% CI) | Monthly decline, % (95% CI) | P-value |
| --- | --- | --- | --- | --- |
| All | Unadjusted | 0.0046 (-0.0044, 0.0135) | -37.0 (-153.4, 26.0) | 0.310 |
| All | Adjusted | 0.0034 (-0.0054, 0.0121) | -26.1 (-130.1, 30.9) | 0.443 |
| ≥18 years | Unadjusted | 0.0036 (-0.0071, 0.0143) | -28.4 (-169.4, 38.8) | 0.500 |
| ≥18 years | Adjusted | 0.0003 (-0.0099, 0.0106) | -2.2 (-107.8, 49.7) | 0.950 |

